# Open Access Data Repository and Common Data Model for Pulse Oximeter Performance Data

**DOI:** 10.1101/2024.08.30.24312744

**Authors:** Nicholas Fong, Michael S. Lipnick, Ella Behnke, Yu Chou, Seif Elmankabadi, Lily Ortiz, Christopher S. Almond, Isabella Auchus, Garrett W. Burnett, Ronald Bisegerwa, Desireé R Conrad, Carolyn M. Hendrickson, Shubhada Hooli, Robert Kopotic, Gregory Leeb, Daniel Martin, Eric D. McCollum, Ellis P. Monk, Kelvin L. Moore, Leonid Shmuylovich, J. Brady Scott, An-Kwok Ian Wong, Tianyue Zhou, Romain Pirracchio, Philip E. Bickler, John Feiner, Tyler Law

**Affiliations:** Department of Anesthesia and Perioperative Care, Zuckerberg San Francisco General Hospital and Trauma Center, University of California San Francisco; University of California San Francisco Hypoxia Research Laboratory; University of California San Francisco School of Medicine; University of California San Francisco Center for Health Equity in Surgery and Anesthesia; Department of Pediatrics, Stanford University School of Medicine; Department of Anesthesiology, Perioperative & Pain Medicine Icahn School of Medicine, New York NY USA; Department of Medicine and Pediatrics, Divisions of Cardiovascular Medicine and Pediatric Cardiology, Stanford University School of Medicine; Department of Medicine, Division of Pulmonary and Critical Care Medicine, University of California San Francisco; Department of Pediatrics, Division of Emergency Medicine, Baylor College of Medicine, Houston, United States; ISO Oximeters Joint Working Group Co-convener; Peninsula Medical School, University of Plymouth, Plymouth, UK; Global Program in Pediatric Respiratory Sciences, Eudowood Division of Pediatric Respiratory Sciences, School of Medicine, Johns Hopkins University, Baltimore, United States; Department of Sociology, Harvard University; Department of Medicine, Division of Dermatology Washington University in Saint Louis; Division of Respiratory Care, Department of Cardiopulmonary Sciences, Rush University; Duke University Department of Medicine, Division of Pulmonary, Allergy, and Critical Care Medicine; Duke University Department of Biostatistics and Bioinformatics, Division of Translational Biomedical Informatics; Division of Biostatistics, University of California, Berkeley

## Abstract

The OpenOximetry Repository is a structured database storing clinical and lab pulse oximetry data, serving as a centralized repository and data model for pulse oximetry initiatives. It supports measurements of arterial oxygen saturation (SaO2) by arterial blood gas co-oximetry and pulse oximetry (SpO2), alongside processed and unprocessed photoplethysmography (PPG) data and other metadata. This includes skin color measurements, finger diameter, vital signs (e.g., arterial blood pressure, end-tidal carbon dioxide), and arterial blood gas parameters (e.g., acid-base balance, hemoglobin concentration).

Data contributions are encouraged. All data, from desaturation studies to clinical trials, are collected prospectively to ensure accuracy. A common data model and standardized protocols for consistent archival and interpretation ensure consistent data archival and interpretation. The dataset aims to facilitate research on pulse oximeter performance across diverse human characteristics, addressing performance issues and promoting accurate pulse oximeters.

The initial release includes controlled lab desaturation studies (CLDS), with ongoing updates planned as further data from clinical trials and CLDS become available.

## Background & Summary

Pulse oximetry is an essential healthcare tool for non-invasively measuring hemoglobin oxygen saturation (SpO2). It is widely used by healthcare professionals to treat, diagnose, and monitor the health of patients worldwide. Despite its critical role, pulse oximeter performance and accuracy are impacted by many factors, though the complete list of these factors and the magnitude of effects need to be better characterized. For instance, it is known that some pulse oximeters may report values higher than the actual arterial functional saturation (SaO2) in patients with darker skin tones, with potential impacts on health care and outcomes.^1–10^ This growing literature highlighting health inequities has triggered a surge of new pulse oximetry research, as well as efforts by regulatory bodies to revise guidance and standards for pulse oximeter testing and performance.^11^

A significant barrier to the success of these research and regulatory initiatives is the lack of sufficient, high-quality data in the public domain. Most publicly accessible data on pulse oximeter performance come from two types of studies: (1) Controlled laboratory desaturation studies in healthy adults and (2) retrospective, observational clinical studies in real-world patient populations. Each source of data has potential limitations, which we hope to overcome with this data repository.

### Controlled Laboratory Desaturation Studies

In a controlled desaturation study, healthy adults undergo precise titration of inspired oxygen to achieve stable arterial oxygen (SaO2) saturation plateaus ranging from ∼70-100%, as measured by arterial co-oximetry measured from radial arterial blood samples.^12^ In a controlled laboratory setting, the data used for comparison of oximeter performance consists of precisely paired and stable SpO2 and SaO2 values. Though there are many other confounding influences, controlled lab studies establish a performance baseline, recognizing that real-world performance will vary. Globally, few laboratories exist that conduct these studies, and even fewer are independent and share data publicly.

### Clinical Studies

Clinical studies in real-world populations include patient populations that are not enrolled in laboratory studies, such as children and neonates, as well as patients with comorbidities and pathophysiologies that impact pulse oximeter performance. These studies provide the opportunity to assess the relationship between device performance, clinical care, and outcomes.^4,9,13^ However, prospective studies are expensive and time intensive,^14,15^ and as a result, most clinical studies to date have been either large retrospective studies or small prospective studies.^16^

Large retrospective studies, where SpO2 and SaO2 are retrospectively paired, usually via an electronic medical record (EMR), can be problematic. Limitations include the inability to ensure proper pulse oximeter probe placement, unknown SpO2 probe type, inability to account for SpO2 signal quality, timing mismatch between SpO2 and SaO2 sampling (i.e. SpO2 and SaO2 may not be from the same time), timing mismatch between SpO2 and SaO2 reporting (i.e. time a sample is drawn and time a sample is recorded in the EMR may not be the same or accurate), and clinical instability (i.e. SpO2 and SaO2 may be dynamic around the time of sample draw due to change in O2 delivery, suctioning or clinical improvement/deterioration). Additionally, data is rarely obtained over the full saturation range of 70-100% as very few patients have stable SpO2 values in the 70% to 88% range without some sort of corrective or dynamic intervention being performed (the exception being patients with congenital cardiac conditions).^17^ Taken together, these factors decrease the reliability and precision of the data needed to characterize the performance of pulse oximeters and their errors. Prospective studies produce higher-quality data and mitigate confounding but they are costly and time-consuming.^18^

The OpenOximetry dataset, introduced in this article, is built to address these challenges, combining data from controlled laboratory desaturation studies with data from multiple prospective studies to create a curated, clean, and reliable open-access source of data for investigators to answer questions regarding the impact of patient variables on pulse oximeter performance, across devices, over time and in different global settings. Secondly, we describe a common data structure for pulse oximeter data sets, including those from controlled laboratory desaturation studies and prospective clinical studies. Future releases of this dataset will include additional pulse oximeter performance tests, clinical trial data, prospectively collected observational data, and other data contributed by other researchers and institutions.

This initial dataset is derived from the Hypoxia Lab^19^ at the University of California, San Francisco (UCSF IRB #21-35637). The repository is intended to serve as a hub for laboratory-based desaturation studies and data from real-world prospective clinical trials, compiling data from ongoing prospective studies generating paired SpO2-SaO2 data that follow similar protocols.^14^ By leveraging a common data structure, otherwise disparate data sets can be harmonized to help answer important questions on pulse oximetry.

## Methods

### Data Acquisition

Data collected to create this dataset was originally based on the dataset requirements for ISO 80601-2-61:2017 and 2013 FDA 510(k) guidelines for pulse oximeter premarket notifications. These initial parameters were expanded to include more parameters known to or hypothesized to impact pulse oximeter performance. The final list of parameters was generated based on discussions via the OpenOximetry Collaborative Community and the EquiOx prospective clinical trial (ClinicalTrials.gov #NCT05554510, Figure 1).

**Figure 1:**
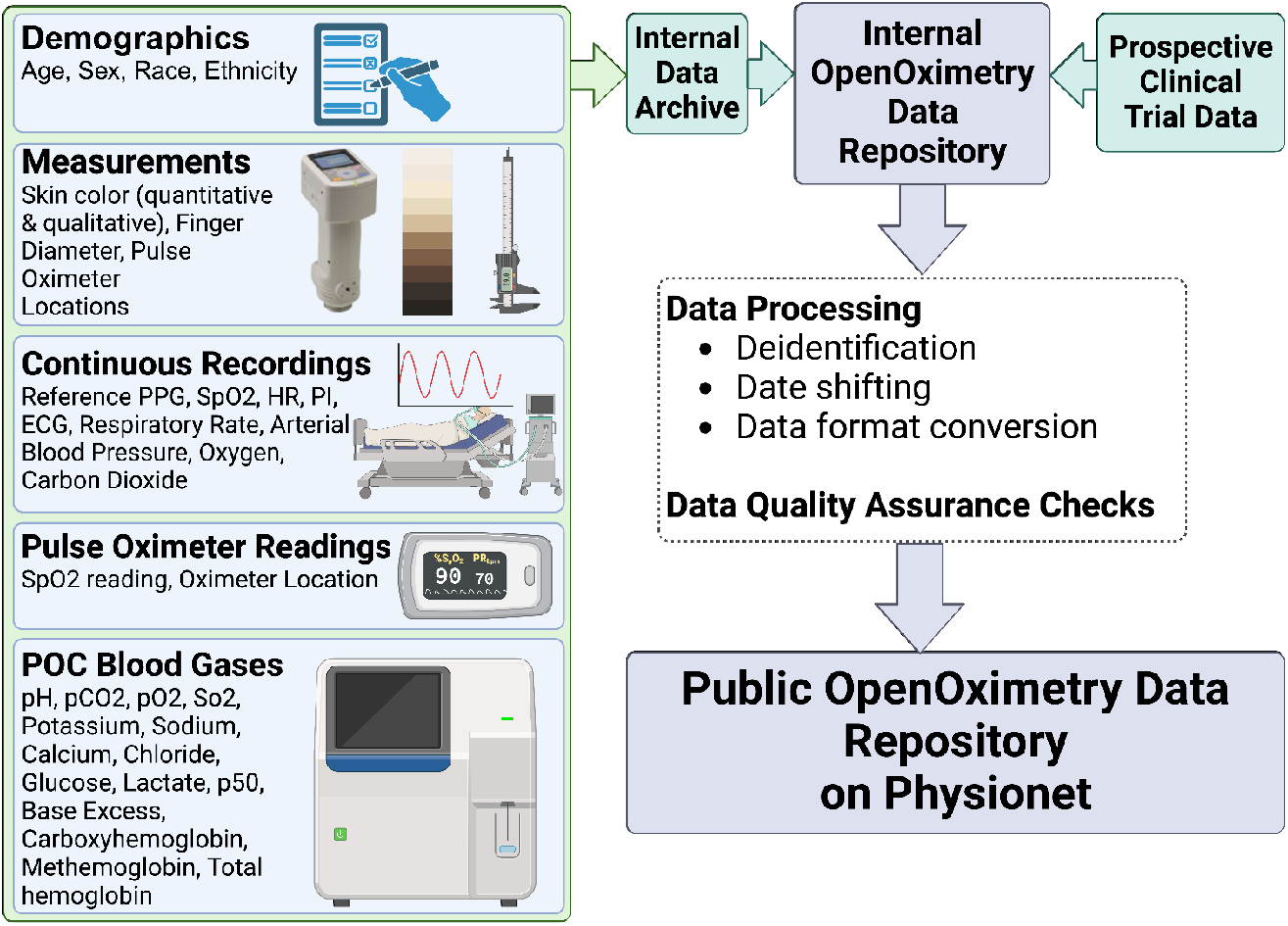
Overview of the OpenOximitry Pulse Oximeter Performance Data Repository. Data are collected from multiple sources at study time and are de-identified, dates shifted, and converted into open and accessible formats before being included in the Data Repository.

Data in the initial database release were collected from healthy humans undergoing controlled desaturation studies that abide by the study protocol from the University of California, San Francisco’s Hypoxia Lab or prospective studies in real-world clinical settings. During a typical controlled desaturation encounter, demographic information, digit diameter, and skin color information are measured and recorded. Radial arterial access is obtained. Test oximeters are placed on the participant and shielded from each other and ambient light. The participant breathes via a closed-circuit system whereby levels of medical air, nitrogen, and carbon dioxide are adjusted. The blend of air and nitrogen is titrated to desaturate the participant to target stable oxygen saturation plateaus distributed over the range of SaO2 70-100% in accordance with ISO80601 standards. These plateaus are estimated using an estimated calculated SaO2;^20^ the sequence of plateaus is depicted in Figure 2.

**Figure 2:**
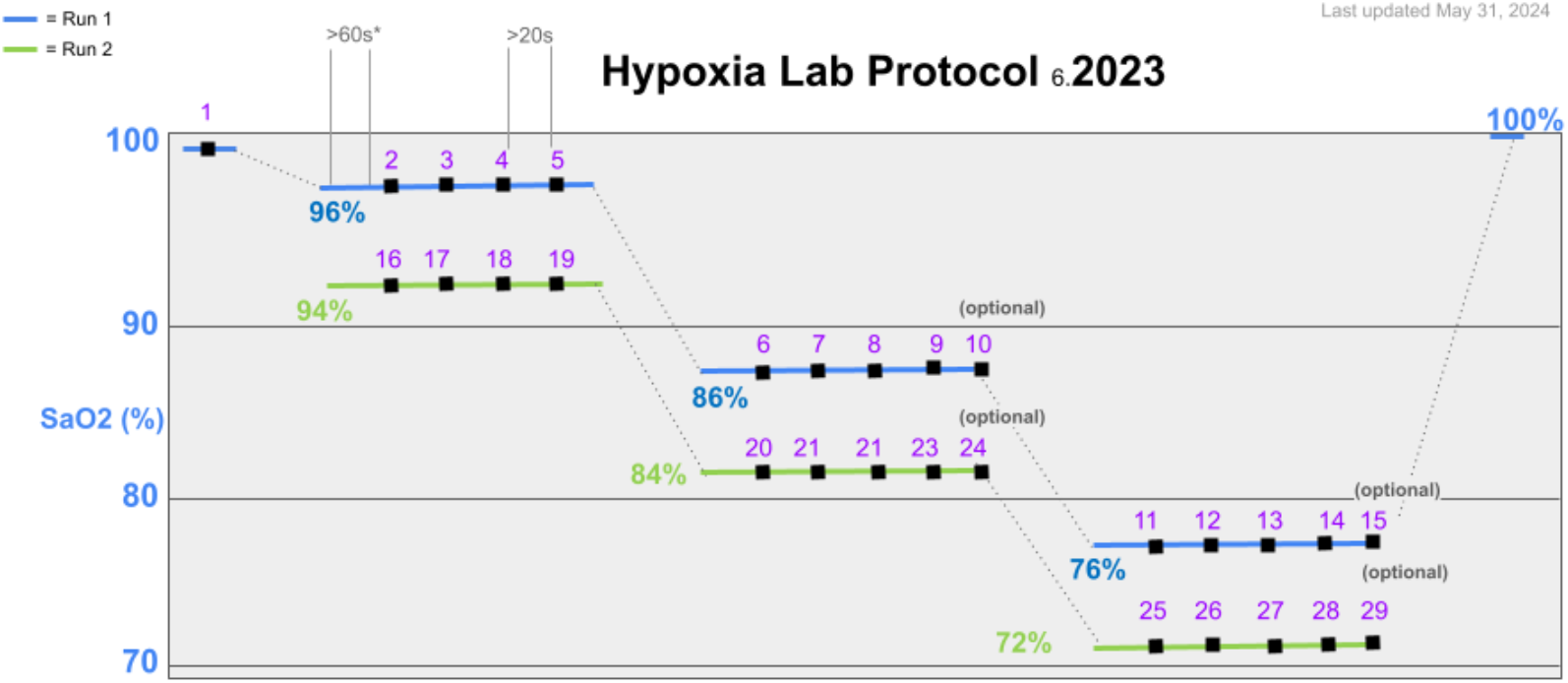
Graphical depiction of the 2023 Hypoxia Lab Study Protocol. Patients are desaturated to the specified SaO2 values, and blood gas samples are drawn while pulse oximeter readings are simultaneously recorded, with each sample/recording being labeled according to the sample number (in purple).

When a target plateau stability is reached, a series of arterial blood gas samples are obtained and immediately run on point-of-care arterial blood gas machines. The saturation reading of each test device at the time the sample is taken is simultaneously recorded and is denoted incrementally as the “sample number” within an encounter. This allows direct comparison between the saturation reading of test devices and the values obtained from point-of-care blood gas machines.

For data captured in prospective, real-world clinical studies, adult or pediatric patients are enrolled in accordance with methods approved by local Institutional Review Boards (IRBs). Methods may vary in accordance with their respective study design but will include measurement of clinical and study pulse oximeter values and arterial blood gas SaO2 values (and the time interval between observations, when applicable).^14^

### De-identification

Study data is de-identified by removing all data considered to be an identifier by the HIPAA safe harbor rules. For each study participant, dates are uniformly skewed by a random number of years into the future, plus (or minus) a randomly assigned number of days to preserve seasonality (e.g. to account for increased or decreased sun exposure during certain times of the year) while obfuscating the exact date the study took place. All ages were less than 89.

Participant and encounter IDs are de-identified by hashing the concatenation of the internal participant identifier and a unique, randomly generated salt for each participant. For example, the string “Participant10_63653” might be hashed for a participant, where “Participant10” is the internal identifier and 63653 is a unique, randomly generated value. This ensures that the chronology of encounters or patients cannot be determined using a rainbow table attack (i.e. prevents someone from generating hashes for “Participant1,” “Participant2,” etc. and creating a lookup table to reverse the hash). Furthermore, tables are sorted by encounter or participant ID so that no chronology can be inferred from the ordering of the flat CSV files.

Only data collected under ethical principles outlined in the Declaration of Helsinki is and will be included in data repository releases.

## Data Records

### Data Tables

The OpenOximetry Repository is a relational database consisting of several tables linked by common identifiers. Most tables have the primary key, encounter_id.

Data tables are intentionally designed to allow other research groups to contribute their data, even from studies that do not capture all of the variables recorded as part of a laboratory-controlled desaturation session. For example, all blood gas data in the database is from arterial samples, but the database also supports the recording of venous blood gas samples. Additionally, data tables are designed such that clinical trials and prospectively captured data can be included in future releases of the dataset.

In selecting the data model for our dataset, we opted against utilizing existing data models, such as Observational Medical Outcomes Partnership (OMOP), for several reasons. While OMOP offers robust capabilities tailored for EMRs and large-scale observational health databases, fundamental components of our dataset are beyond the scope of an EMR-centric data model. For example, objective and subjective measures of skin color, finger measurements, and pulse oximeter device data do not easily fit into the OMOP data model. Additionally, we prioritized accessibility for non-technical users to contribute and navigate the data effectively. Given the complexity of OMOP’s schema and its emphasis on structured clinical data, we deemed a simpler data model more suitable for our multi-source dataset, facilitating ease of integration and interpretation across a wider range of contributors. The OMOP common data model has 37 tables and 394 fields, while our data model has six tables and 138 fields. This decision was based on stakeholder input via the Open Oximetry Project Collaborative Community and aligns with our goal of fostering inclusivity and collaboration within our research community, ensuring that all stakeholders can engage meaningfully with the data.

A general description of the tables is given in Table 1 and as follows:

**Table 1:**
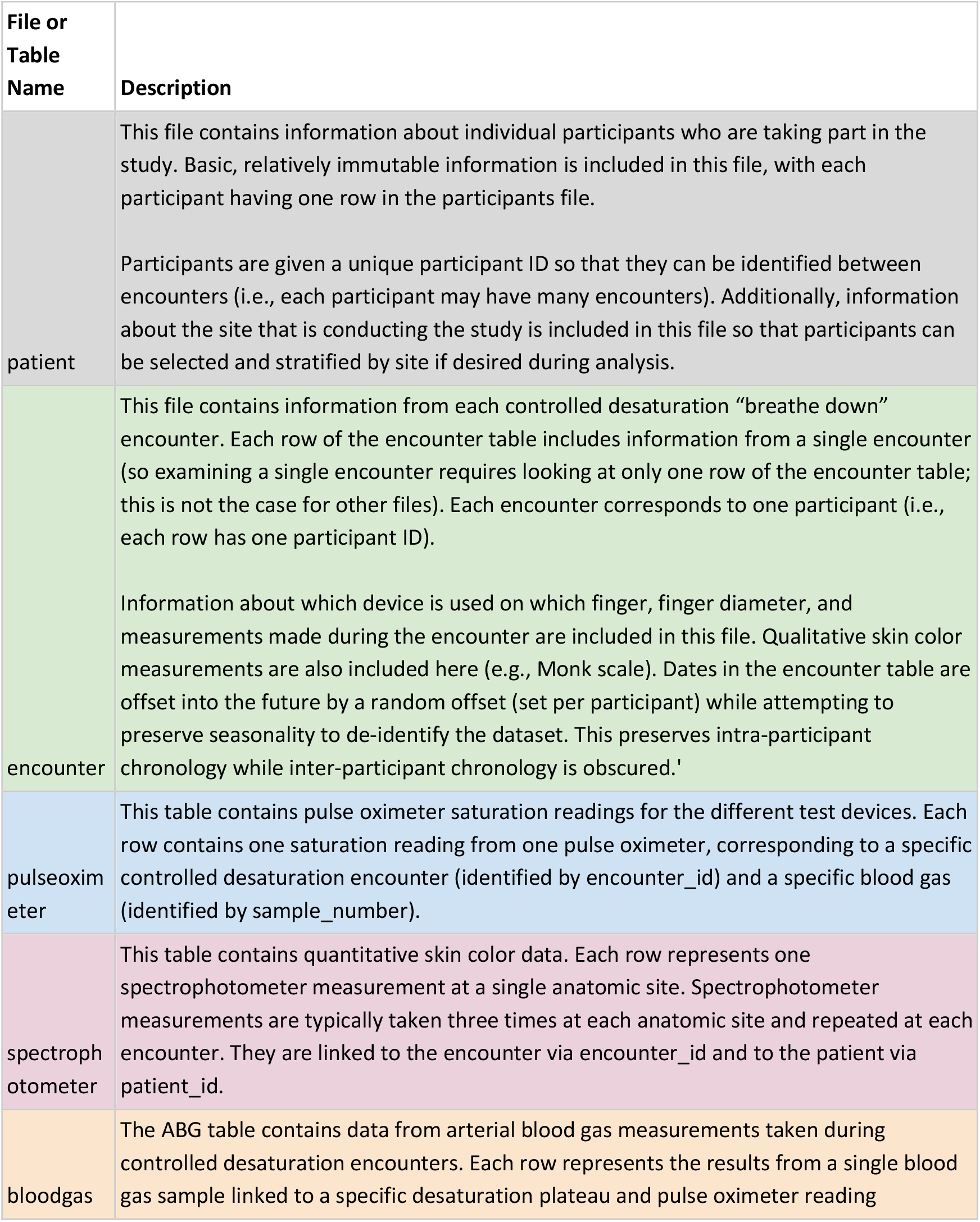
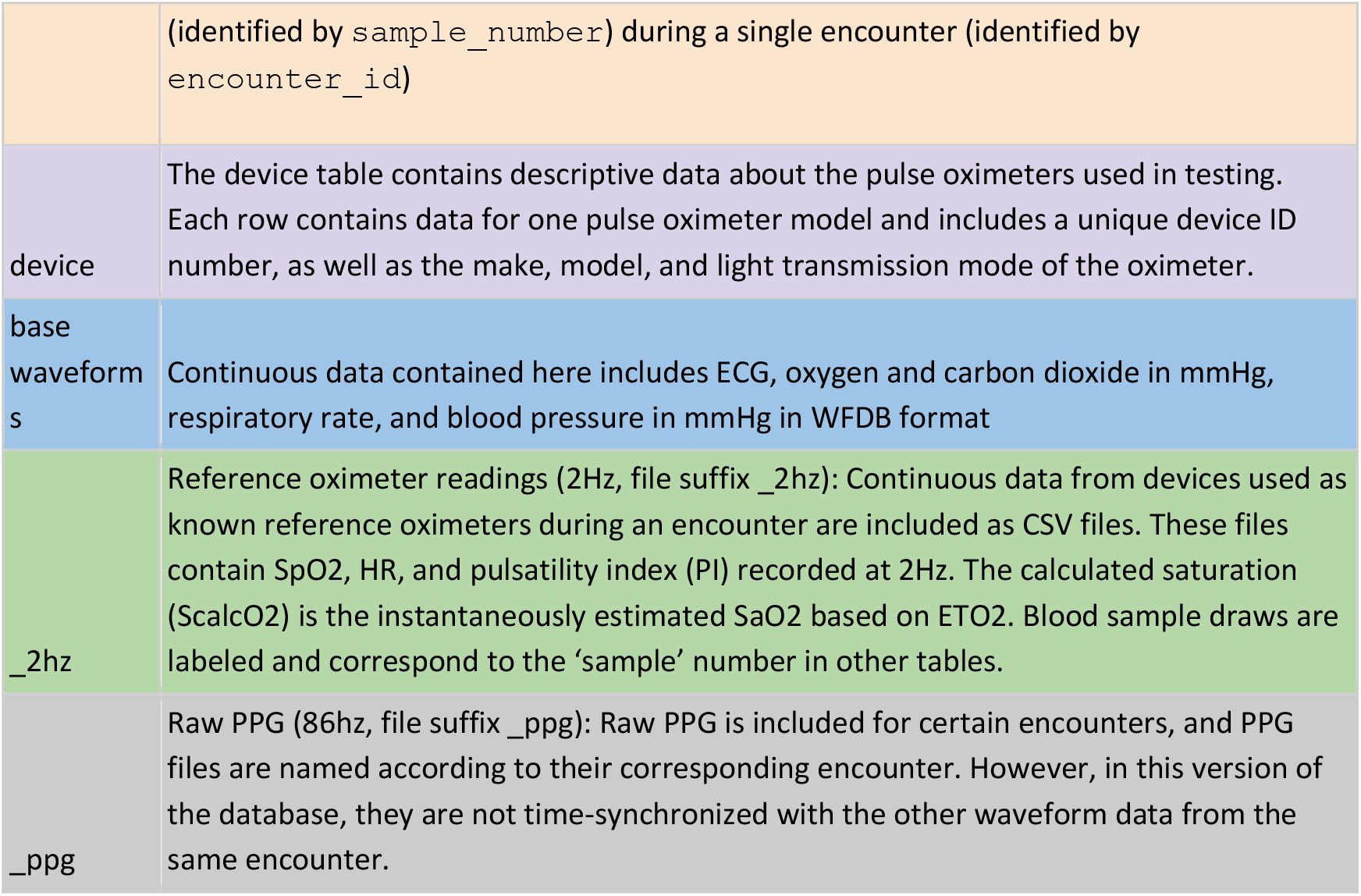
Description of each table or file suffix in the data repository.

#### Patients

This file contains information about individual patients/participants that are taking part in the study. Basic, relatively immutable information, like the patient’s sex, ethnicity, and racial identity, are included in this file, with each participant having one row in the participants file. Participants are given a unique participant ID so that they can be tracked between encounters (i.e., each participant may have many encounters). Additionally, information about the study site is included so that participants can be selected and stratified by site if desired during analysis.

#### Encounter

This file contains information from each controlled desaturation encounter (or, if prospective data, hospital event). Each row of the encounter table includes information from a single encounter. Examining a single encounter requires looking at only one row of the encounter table; this is not the case for other files. Each encounter corresponds to one participant (i.e., each row has one participant ID). Information about the participant’s current age, which device is used on which finger, finger diameter, and qualitative skin color measurements (e.g., Monk scale at multiple anatomic sites) are included in this file. Dates in the encounter table are offset into the future by a random offset (set per participant) while attempting to preserve seasonality to de-identify the dataset. This preserves intra-participant chronology while inter-participant chronology is obscured.

#### Pulse oximeter

This table contains pulse oximeter saturation (SpO2) readings for the different test devices. Each row contains one saturation reading from one pulse oximeter, corresponding to a specific controlled desaturation encounter (identified by encounter_id) and a specific blood gas (identified by sample_number). Sample numbers refer to the purple numbers corresponding to blood gas samples taken, as depicted in Figure 1.

#### Blood Gas Co-oximetry

The blood gas table contains blood gas data, including arterial functional saturation (SaO2). Each row represents the results from a single blood gas sample and clinical data from that time. These samples are linked to pulse oximeter readings and other data during the same encounter via ‘sample_number’ and ‘encounter_id’. As in the pulse oximeter table, sample numbers from controlled desaturation studies refer to the purple numbers corresponding to blood gas samples taken, as depicted in Figure 1.

#### Device

The device table contains descriptive data about the pulse oximeters used in testing. These data will be published when available, though they may not always be available and are not required for publishing data in this repository. Each row contains data for one pulse oximeter model and includes a unique device ID number, as well as the make, model, and light transmission mode of the oximeter (and probe).

#### Spectrophotometer

This table contains quantitative skin color and reflectance data. Each row represents one spectrophotometer measurement at a single anatomic site. Spectrophotometer measurements are typically taken three times at each anatomic site to ensure data consistency and are repeated at each encounter. They are linked to the encounter via encounter_id and to the patient via patient_id. Further information can be found on the Hypoxia lab skin color assessment protocol page.^21^

### Waveform Files

Continuous waveform data is stored in WFDB or CSV format, with file names corresponding to the encounter.

Up to three different waveform files are available for each encounter:

- (150hz, base file name): Continuous data contained here includes ECG, oxygen, and carbon dioxide in mmHg, respiratory rate, and blood pressure in mmHg. Recordings are made to 3 decimal places.
- Reference oximeter readings (2Hz, file suffix _2hz): Continuous data from devices used as known reference oximeters during an encounter are included as CSV files. These files contain SpO2, HR, and a signal quality indicator recorded at 2Hz. The calculated saturation (ScalcO2) is the instantaneously estimated SaO2 based on ETO2. Blood sample draws are labeled and correspond to the ‘sample’ number in other tables.
- Raw PPG (86hz, file suffix _ppg): Raw PPG is included for certain encounters, and PPG files are named according to their corresponding encounter. Both infrared and red data are stored, and this data is both unscaled and unfiltered. However, in this version of the database, they are not time-synchronized to the other waveform data from the same encounter.

## Technical Validation

Data changes from time of storage to repository upload were minimized as much as possible. Most processing was done to de-identify data, convert waveform files to WFDB format, and validate de-identification and data conversion. Waveform data was stored in plain text format during studies and was converted to WFDB format without further processing.

### Data storage

The raw data is stored in a series of databases on REDCap (Vanderbilt University, Nashville, TN, USA), a web-based tool designed to capture data for clinical research. REDCap was chosen for a number of reasons: (1) it allowed for structured data storage similar to traditional databases, (2) it has a graphical front-end that allows for manual data entry of database fields (e.g., SpO2) by research staff, (3) data can be updated and accessed programmatically via the API, and (4) REDCap is FDA compliant with 21 CFR Part 11. The raw data generated by blood gas analyzers, lab equipment, and research staff are ingested by a series of Python scripts to generate the public-facing version of the database.

The code used to build the database is stored under version control and was regularly reviewed by laboratory members. Frequent feedback from colleagues, as well as embedded code to assert and verify that the data that was being saved in the repository matches the data that was saved at study time, helped further validate the data stored in the respiratory. For example, the code used to build the database asserts that one of the waveform files written to disk matches the waveform data that was intended to be stored by reading the file from disk and comparing the values. If the values do not match, the data processing code will cause an exception, alerting whoever is running the code of the inconsistency. The repository build process generates graphs of critical measured parameters, allowing for verification that the dataset contains rational values. Additionally, some data in flat files were manually verified against the data stored at the time of the study to ensure accuracy.

## Usage Notes

### Data Access

The OpenOximetry database contains information about research participants and the care of patients, so access is restricted. Researchers are required to request access to the database formally. Access can be requested through Physionet^22^ by making a free account and agreeing to the data use agreement.

### Joining Tables

Users may filter and join tables to obtain the data set required. For instance, to examine the paired SpO2 and SaO2 measurements for a specific pulse oximeter, the user would first filter the ‘encounter’ table based on ‘device_id’. This table would then be joined on ‘encounter_id’ and ‘device_id’ to the ‘pulseoximeter’ table (to obtain SpO2 values) and to the ‘abg’ table on ‘encounter_id’ and ‘sample’ (to obtain the ABG results for each sample during the encounter). Demographics or skin color data could be added by joining again on the ‘patient’ or ‘spectrophotometer’ tables.

High-resolution waveform files included in this repository are stored in WFDB format. Within the waveform directory, intermediate directories 0-9 and a-f contain all waveform records for encounters that begin with that character. For example, the waveform records for encounter b94d27b9934d3e08a52e52d7da7dabfac484efe37a5380ee9088f7ace2efcde9 would be found in the b intermediate directory, as the first character of the encounter ID is b. Additionally, raw PPG data in this release is available for only a subset of encounters as a separate, non-time-synced WFDB file (suffix _ppg), recorded at 86 Hz.

Agreeing to the terms of a Data Use Agreement is required to access the data. Terms of this agreement include but are not limited to not attempting to identify any individual in the dataset, maintaining dataset security, not selling or distributing data, and using the data in the dataset only for scientific research without resale of the data.

## Data Availability

All data produced are available online at Physionet

https://physionet.org/content/openox-repo/

## Acknowledgments

The Open Oximetry Project has been supported by the Gordon and Betty Moore Foundation, The Patrick J. McGovern Foundation, the Robert Wood Johnson Foundation, Unitaid through PATH, and USAID STAR.

We would additionally like to extend our sincere gratitude to Odi Ehie, Elizabeth Namugaya Igaga, and Sandy Weininger for their valuable contributions to this project.

AIW is supported by REACH Equity under the National Institute on Minority Health and Health Disparities (NIMHD) of the National Institutes of Health under U54MD012530.

GWB is a recipient of the Medtronic Research Award. Medtronic was not involved in study design or any portion of the study.

SH is supported by the National Heart, Lung, Blood Institute under K23HL169901.

EPM is supported by DP2MH132941.

## Author Contributions

Conceptualization: NF, MSL, TL

Data collection: EB, YC, SE, LO

Repository and data model architecture, build, and deployment: NF, TL

Data analysis, data interpretation: NF, TL, RK, TZ

Writing — original draft: NF, TL, MSL

Writing — review and editing: NF, TL, MSL, EB, YC, SE, RK, LO, PB, JBS, EDM, SH, AIW, GWB, EM, CSA, DRC

## Code Availability

A Python library of useful functions for analysis of the OpenOximetry dataset is available for use upon request. The code used to generate the dataset is also available upon reasonable request; however, some aspects of this code (e.g., API keys, random number generator seeds) may be obfuscated to prevent unauthorized access of data or re-identification of patients or comply with privacy laws and regulations.

